# The virtual multiple sclerosis patient: on the clinical-radiological paradox

**DOI:** 10.1101/2023.12.01.23299274

**Authors:** P Sorrentino, A Pathak, A Ziaeemehr, E Troisi Lopez, L Cipriano, A Romano, M Sparaco, M Quarantelli, A Banerjee, G Sorrentino, M Hashemi, V Jirsa

## Abstract

Multiple sclerosis (MS) is typically diagnosed based on the clinical presentation, the presence of structural MRI lesions, and a “no better explanation” criterion. The structural lesions, disseminated in time and space, are a consequence of autoimmune processes leading to the damage of the myelin sheath in the central nervous system. As such, one would expect that more lesions would relate to higher clinical disability. However, a conflicting scenario is often present, with a high lesion load related to mild clinical impairment, and vice versa, a phenomenon referred to as the “clinico-radiological paradox”. The myelin damage in MS is widespread, which is likely mirrored in a widespread slowing of conduction velocities. However, conduction velocities are typically measured on selected white-matter tracts (e.g., visual evoked potentials), which do not directly relate to clinical impairment. In this paper, we hypothesize that the overall slowing of conduction velocities (i.e., across all brain tracts) is a better predictor of clinical disability. However, estimating the whole-brain average velocities is challenging. To overcome this obstacle, we estimated patient-specific conduction velocities in MS patients by merging multimodal data (i.e., DTI and source-reconstructed magnetoencephalography) to inform large-scale brain models, fitted on each individual patient. We started from the known reduction of the power of the alpha frequency band, as well as the shift in its peak, observed in MS patients. We then reproduced these individual spectral features in silico using large-scale models based on the individual connectomes. We then used state-of-the-art deep neural networks for Bayesian model inversion to estimate the most likely average conduction velocity in each patient, given the observed spectral features (and the connectomes). Finally, we used the inferred conduction velocities to predict the individual clinical disability. We find that the conduction velocities inferred for patients are significantly lower than those inferred for controls and that they are predictive of individual clinical disability, well above the predictive power of demographic and clinical variables and lesion load. Our results suggest a biologically and physically plausible solution to the “clinico-radiological” paradox, where the inferred, individual changes in conduction velocities across the whole networks are proposed as causative to the clinical disability.

## Introduction

Multiple sclerosis (MS) is an inflammatory disease of the human central nervous system^1^. While the aethiology of the disease remains unknown to this day, structural magnetic resonance has added to our understanding by allowing in-vivo, non-invasive quantification of damage.^2^ The underlying assumption here was that patients with a greater lesion load would be clinically worse off than those with a smaller lesion load, and patients with more new lesions would be clinically more compromised than those without as many new lesions.^3^ However, converging evidence showed otherwise, proving that lesion load alone remains a poor predictor of clinical outcome.^4–7^ This phenomenon has been referred to as the “clinico-radiological paradox”, which makes it more difficult to utilize the lesion load in clinical practice and even as a biomarker in clinical trials. This explanatory gap points to the existence of more complex mechanisms, i.e., ’hidden variables’, that mediate the causal relationship between structural damage and clinical disability in MS. Multiple potential mechanisms (i.e., ‘Candidate variables’) and explanations of the clinico-radiological paradox have been attempted, but failed to provide a convincing, overarching understanding of the phenomenon.^3^

In the context of systems neuroscience, the brain is typically conceptualized as a complex dynamical network, whereby the accurate causal prediction on the effect(s) of a perturbation (e.g., a lesion) is challenging due to nonlinear interactions between functional units and consequently, the absence of one-to-one correspondence between structure and function.^8–10^ Hence, using such prism one would not expect the lesion load to directly predict the final behavior outcome (i.e., the clinical condition). In particular, lesions in MS (especially in the early phases, and with the exception of the primary progressive forms) are believed to relate to attacks from the immune system to the myelin sheath, which in turn manifest themselves as slower conduction velocities^11^ (i.e., higher delays in the propagation of impulses traveling across the white-matter bundles). Recent evidence confirmed that there are widespread augmented delays across the brain in MS patients as compared to healthy controls.^12^ Importantly, there is a wide body of literature in statistical mechanics that is concerned with the behavior of complex networks as a function of the delays.^13,14^ As such, one can estimate analytically and/or numerically how augmented delays would affect the overall network organization (specifically in terms of frequencies of network oscillations).^15–17^ In particular, we focus on the fact that a shift in the power spectra has been observed in MS, whereby patients with MS show lower power in the alpha frequency band (8-13 Hz) and a shift in the peak frequency toward slower frequencies.^18–20^ Even during tasks, alpha rhythms are most prominently affected in MS.^21^

In this paper, we hypothesize that the delays that are induced by the damage to the myelin in MS are responsible for the observed specific changes in power spectrum, which in turn would be better predictors of clinical disability (as opposed to the total lesion load *per se*). To test this hypothesis, we utilized a previously published dataset^12^, made of 18 patients with MS and 20 controls. All subjects underwent both a magnetoencephalography scan and an MRI scan, which included the estimation of the tractography. This way, we have access to the individual connectomes and to the large-scale activity (i.e., the source-reconstructed MEG signals) for each subject. Firstly, we tested in this dataset for the presence of the previously described shift in the alpha-frequency peak and the reduction of the alpha-band in the power-spectra in MS patients. Once this was confirmed, we used large-scale mechanistic brain modeling^16^ to show that, given the individual structural connectomes, such changes on the overall power-spectra could be theoretically expected by higher conduction velocities.^22^ The large-scale brain network modeling approach emphasizes the whole-network character of the changes in the organization of brain activities, which provides a link between the underlying physiological mechanisms and the clinical disability.^23^ Then, once such theoretical prediction was confirmed, we moved on to test if the subject-specific reduction of conduction velocities could be inferred using unsupervised machine-learning techniques, starting from the empirical MEG data and the connectomes.^24^

However, the parameter estimation of large-scale brain network models is a challenging task due to the non-linearity of the dynamics at each brain area and the strength of relationships between variables and/or parameters, as well as to the large dimensionality of the latent space.^25^ To this end, advanced machine learning algorithms designed for high-dimensional probabilistic models are required to efficiently estimate the unknown model parameters and the statistical relationships between variables, ideally including the uncertainty.^26^ In this study, we used Bayesian inference for model inversion, which is a principled method for updating beliefs with the information provided by the observed data (new evidence) to quantify uncertainty over hidden variables. For the present manuscript a key goal would be to infer patient specific average conduction velocities as the parameter that modulates power spectra in the alpha frequency band. To this end, we used simulation-based-inference (SBI)^27^ to estimate the conduction velocities (and a scaling parameter) over the patient’s structural information, with the aid of only forward simulations. To efficiently carry out SBI, we employed state-of-the-art deep neural networks for conditional probability density estimation,^24^ which provide the posterior distribution of the individual delays conditioned on the individual power-spectra in the alpha band. If our hypothesis is correct, we would expect that the model inversion would provide us with longer estimated delays for MS patients as compared to healthy controls. Finally, as explained earlier, we set out to test our last hypothesis, namely that individual delays would be a better predictor of clinical disability as compared to the lesion load alone. To this end, we built a multilinear statistical model to predict individual clinical disability.^28^ Besides other demographic and clinical features, we also included the total lesion load as a predictor. Then, we added the individual average delays that were inferred by the Bayesian model inversion. Given our hypothesis, we expect that including the individual delays should significantly improve the prediction of the individual clinical disability beyond the predictive power of the individual lesion load.

## Materials and methods

### Participants

The participants were recruited at the outpatient clinic of the Institute for Diagnosis and Cure Hermitage Capodimonte (Naples, Italy). MS was diagnosed according to the revised 2017 McDonald criteria,^29^ with exclusion criteria defined as age < 18 years, recent clinical relapse and/or steroid therapy (i.e., 3 months before the study), use of illicit drugs, stimulants, amphetamines, barbiturates, and cannabinoids; history of central nervous system disorders other than MS, severe mental illness, inability to understand and complete “patient reported outcomes” and cognitive evaluation, or inability to undergo the MRI scan. All patients underwent a neurological clinical examination, Expanded Disability Status Scale (EDSS) scoring, the Symbol Digit Modalities Test (SDMT) to measure cognitive impairment, the Fatigue Severity Scale (FSS), and the Beck Depression Inventory (BDI) to assess depression. The controls for the MS cohort were selected among the caregivers and spouses of the patients. Genetic relatives were not allowed as controls. The main features of the MS cohort are summarized in Table 1. The study was approved by the local Ethics Committee (Prot.n.93C.E./Reg. n.14-17OSS) and all participants provided written informed consent in accordance with the Declaration of Helsinki.

**Table 1.** Features of the multiple sclerosis cohort.

|  | **CONTROLS** | **MS** | **p-value** |
| --- | --- | --- | --- |
| Age (years) | 45.8 (± 11) | 44.9 (± 9.9) | 0.8 |
| Education (years) | 13.6 (± 3.8) | 13.8 (± 5) | 0.9 |
| Gender (m/f) | 6/14 | 6/12 | 0.3 |
| Disease duration (months) | - | 187.7 (± 131.8) | - |
| EDSS (mean + SD) | - | 4.5 (± 1.9) | - |
| SMDT (mean + SD) | - | 40.3 (± 13) | - |
| FSS (mean + SD) | - | 36.1 (± 14) | - |
| BDI (mean + SD) | - | 12.8 (± 1.3) | - |
| LESION LOAD (mean + SD) | - | 12959 (± 12253) mm<sup>3</sup> | - |

**Abbreviations:** EDSS: Expanded Disability Status Scale; SDMT: Symbol Digit Modalities.
FSS: Fatigue Severity Scale; BDI: Beck Depression Inventory; LL: lesion load.

### MRI acquisition and processing

The data was processed as in ref.^12^ In short, each MRI scan was performed immediately after the MEG recording on the same MRI scanner (1.5 Tesla, Signa, GE Healthcare). Analyzed sequences included echo-planar imaging for DTI reconstruction (TR/TE 12,000/95.5 ms, voxel 0.94×0.94×2.5 mm3, 32 equally spaced diffusion-sensitizing directions, 5 B0 volumes) and 3D-FLAIR volume for WM lesion segmentation (TR/TE/TI 7000/145/1919 ms, echo train length 170, 212 sagittal partitions, voxel size 0.52×0.52×0.80 mm3). Preprocessing of the diffusion MRI data was carried out using the software modules provided in the FMRIB software library (FSL, http://fsl.fmrib.ox.ac.uk/fsl). All diffusion MRI datasets were corrected for head movements and eddy current distortions using the “eddy_correct” routine,^30^ rotating the diffusion sensitizing gradient directions accordingly,^31^ and a brain mask was obtained from the B0 images using the Brain Extraction Tool routine.^32^ A diffusion-tensor model was fitted at each voxel, and fiber tracts were generated over the whole brain by deterministic tractography using the Fiber Assignment by Continuous Tracking (FACT) algorithm implemented in Diffusion Toolkit (angle threshold 45°, spline-filtered, masking by the FA maps thresholded at 0.2). A cortical study-specific Region of Interest (ROI) datasets was obtained by masking the ROIs available in an MNI space-defined volumetric version of the Desikan-Killiany-Tourville (DKT) ROI atlas^33^ using the GM tissue probability map available in SPM (thresholded at 0.2). This was done to limit the endpoints of the fibers to cortical and adjacent juxtacortical white matter voxels in the subsequent ROI-based analysis of the tractography data. To obtain the corresponding patient-specific ROI sets, each participant’s FA volume was spatially normalized^34^ to the FA template provided by FSL using SPM12, and the resulting normalization matrices were inverted and applied to the ROI set.

### MEG pre-processing

MEG pre-processing and source reconstruction were performed as in ref.^35^ Preprocessing and source reconstruction operations were carried using the Fieldtrip Toolbox.^36^ Each participant underwent a MEG recording, composed of both eyes-closed resting-state segments of 3’30” each. Four anatomical coils were applied on the head of each participant and their position was recorded along with the position of four head anatomical points, to identify the position of the head during the recording. Eye blinking (if present) and heart activity were recorded through electro-oculogram (EOG) and electrocardiogram (ECG), to identify physiological artifacts.^37^ An expert rater checked for noisy signals and removed them. An anti-alias filter was applied to the MEG signals, acquired at 1024 Hz, before being filtered with a fourth order Butterworth IIR band-pass filter (0.5-48 Hz). We used principal component analysis^38,39^ and supervised independent component analysis^40^ to remove the environmental noise and the physiological artifacts, respectively.

### Source reconstruction

The source reconstruction was performed as in ref.^12^ In brief, the signal time series were reconstructed using 84 ROIs based on the DKT atlas. The reconstruction took place utilizing the volume conduction model proposed by Nolte.^41^ Based on the native MRIs of each subject, the linearly constrained minimum variance (LCMV) beamformer was applied to reconstruct the signal sources based on the centroids of each ROI.^42^

### Power-spectra estimation

Power spectral density (PSD) for each source-reconstructed time-series was estimated by the Welch method as implemented in MATLAB.^43^ In short, each time-series is divided into the longest possible segments to obtain as close to but not exceed 8 segments with 50% overlap.^43^ Each segment is windowed with a Hamming window. The modified periodograms are then averaged to obtain the PSD estimate.^43^ The resulting PSD estimates were further grouped across ROIs to obtain a median PSD for each participant. The peak alpha frequency, peak alpha spectral density and total alpha power (area under the spectral density curve between 8-13 Hz) were estimated from the resulting median PSD. Differences between groups were tested using the Mann-Whitney U-test, and then Bonferroni corrected across the three parameters explored.

### Numerical solution of the Stuart - Landau model

Each ROI was conceptualized as an autonomous Stuart-Landau oscillator.^16,44,45^ The Stuart-Landau system possesses either damped or limit-cycle solutions, depending on the bifurcation parameter *a*.^45^ For *a* < 0, the system exhibits damped oscillations, akin to a pendulum subject to friction. In response to noise, the system relaxes back to its stable fixed point by executing oscillations specified by angular frequency *ω*_0_. The rate of amplitude damping is dictated by |*a*|. In contrast, limit cycle solutions exist for *a* > 0; in this regime, the system can oscillate in a self-sustained fashion, even in the absence of external noise. Oscillators are connected to each other via white-matter, with coupling strength specified by the (subject-specific) DTI fiber counts (*c_jk_*). Adjacency matrices are scaled by a global coupling parameter (*G*). Coupling between ROIs is subject to finite conduction times, estimated by dividing inter-node euclidean distances by an average conduction velocity, 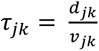. The activity of each ROI is given as:

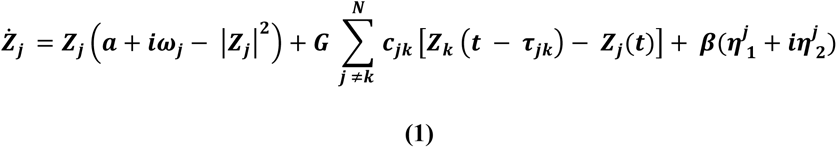

Where *Z* is a complex variable and *Re*[*Z*(*t*)] is the corresponding time-series. Each ROI is regarded as a local, feedback inhibition driven E-I unit, oscillating in the gamma band. Accordingly, the natural frequency of each ROI was set to 40 Hz (*ω_j_* = *ω*_0_ = 2 * π * 40). This study is conducted with *a* = –5, to capture the damped nature of local oscillations. Moreover, *a* = –5 best captures the slowest decay time constants of GABA_B_ inhibitory receptors (∼1s).^16^ Noise, with amplitude *β* = 1 was added to each oscillator to model stochastic fluctuations.

The expected spectral summary statistics were calculated, applying machine learning techniques to large-scale mechanistic models of the brain, given a priori range of parameters. The deep neural network was built to be invertible, meaning that, once trained, it can generate spectral properties from the delays, as well as estimating the delays from the spectral properties. Then, the empirical spectral properties were used as the input to the ’reversed’ algorithms (conveyed by the reversed gradient of colors in the figure), which provided us with the most likely conduction velocity (in each subject). The inferred velocities were then added to a multilinear model, along with demographic, clinical and structural information, in order to predict the individual disability.

Since the object of the study is to model alpha oscillations, we restrict our attention to the regime spanned by *G* = 800 – 2000 and 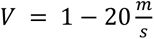 (see Figs 3-5). Simulations were run for 100s by numerically integrating the system of equations (Eq.1), through the Euler-Murayama method. The first 10s were discarded as transients. The PSD was estimated for the remaining 90s, using the *pwelch.m* function. This process was repeated for each subject’s connectome to infer the subject-specific optimal working point.

### Simulation-based inference on whole-brain parameters

Next, we use probabilistic machine learning based on a large-scale brain network model of MEG to infer the individual conduction velocities in MS patients (and controls). In brief, we built a function that, given any particular delay (and coupling parameter), provides us with the expected peak-frequency, peak-amplitude and total power in the alpha band. The key point is that such a function is required to be smooth, invertible. Hence, we then invert this function to estimate the most likely individual velocities (and coupling parameter), given the observed individual spectral-features.

More in detail, the Bayesian approach we use offers a principled method to perform probabilistic inference and prediction, while the uncertainty in the parameter estimation is naturally quantified through probability distributions placed on the parameters updated with the information provided by the data. Formally, given data *y* and model parameters *θ*, Bayes rule defines the posterior distribution as *p*(*θ*|*y*) ∝ *p*(*y*|*θ*) *p*(*θ*), where *p*(*θ*) is the prior probability distribution (over a plausible range of values), and *p*(*y*|*θ*) is the likelihood distribution, which quantifies the probability that the observation *y* is generated by a specific set of *θ*. Here, *θ*={*G*, *V*}, with *G* defined as the global coupling and *V* as the average conduction velocities across the white matter bundles, whereas *y* represents the PSD features (peak frequency, peak amplitude and total power in the alpha band), observed in the MEG data of each subject. The key challenge to efficiently perform Bayesian inference lies in the evaluation of the likelihood function *p*(*y*|*θ*), given the samples from the prior distribution. In other words, one needs the calculation of intractable integrals to evaluate the likelihood of observing *y* given a certain value of *θ* (that is, the delays and coupling). In particular, for inference on MEG data at the whole-brain scale, the computational cost of evaluating the likelihood function becomes prohibitive, rendering the likelihood-based approaches, such as Markov chain Monte Carlo sampling, inefficient in practice.^26^ Simulation-based inference (SBI), or likelihood-free inference, performs efficient Bayesian inference for high-dimensional complex models where the calculation of the likelihood function is either analytically or computationally intractable.^27^ SBI employs deep artificial neural networks (ANNs) to learn an invertible transformation between parameters and data features from a budget of simulations performed with parameters randomly drawn from the prior distribution (as opposed to sampling from the prior distribution and then explicitly evaluating the likelihood function). Using this approach, a simple baseline probability distribution (e.g., a uniform or standard normal distribution) is converted into a more complex distribution (i.e., posterior distribution), through a sequence of invertible transformations (implemented via deep neural networks). In this context, density estimators reconstruct the probability density function (i.e., the relationship between the outcomes of a random variable and its probability) using a set of given data points, without having access to all the possible outcomes. Traditional density estimators such as histograms and kernel density estimators typically perform well only for a small number of random variables. In contrast, recently advanced neural density estimators based on ANNs scale better with the number of random variables, and can incorporate domain knowledge in their design.^46^ In this study, we used a Masked Autoregressive Flow (MAF),^47^ which supports invertible nonlinear transformations, and enables highly expressive transformations with low-cost computation.

In our case, to perform SBI, three inputs need to be specified: (i) a prior distribution describing the possible conduction velocities (and global coupling), (ii) a mechanistic model that simulates the large-scale activity in case of any particular conduction velocity and global coupling (here, the Stuart-Landau model coupled according to the individual connectomes) and, (iii) a set of observed MEG data (or low-dimensional data features) as the target of the fitting (in this context, the observed spectral features). We trained a MAF with a budget of simulations given random parameters drawn from the priors. After the training step, one can quickly evaluate the posterior distribution for arbitrary new observations or empirical data by a forward pass through the trained MAF. By providing fast model simulations and extracting informative but low-dimensional data features (such as the PSD), this methodology efficiently provides the posterior distribution of model parameters at the whole-brain scale,^26^ since it requires neither model nor data features to be differentiable.^24^

After the training step and the posterior estimation, we proceeded to the sensitivity analysis by calculating the eigenvalues and the corresponding eigenvectors.^24^ A strong eigenvalue indicates that the gradient of the posterior is large and, hence, the system output is sensitive to changes along the direction of the corresponding eigenvector. This will allow us to compare the relative importance of the two inferred parameters (i.e., the velocities and the global coupling) on the outcome measures (i.e., the observed spectral properties).

In this study, the SBI on the whole-brain model of Stuart-Landau oscillators is first validated using synthetic data generated with the subject-specific structural connectomes, as to recover the ground-truth parameters, and then applied to empirical MEG data. To this end, we used the PyTorch-based SBI package,^48^ and a simulation budget size of 20000 simulations in order to train the MAF. The parameters were drawn randomly from a uniform prior in the ranges: *G* ∈ [800, 2000], and *V* ∈ [1, 20]. By transforming the observation from the time-domain to the frequency-domain,^49^ we utilized the summary statistics of PSD (amplitude, median frequency, and the total alpha power) as the data feature for the training and the inference steps. This was done, as explained earlier, based on the relationship we demonstrated to exist between the delays of Stuart-Landau oscillators (weakly coupled according to the patient’s connectome), and the frequency of oscillations across the network. The model simulation and parameter estimation were performed on a Linux machine with Intel Core i9-10900 CPU 2.80GHz and 32 GB of RAM.

### Prediction of clinical impairment via a multimodal linear model

We then moved on to test the hypothesis that the inferred delays could improve the prediction of the individual clinical impairment as compared to only including the total volume load in the whole-brain model. To test this hypothesis, we built a multilinear model to predict the individual clinical disability (as estimated by the EDSS) based on the demographics, the clinical data and the total lesion load. Specifically, we considered the EDSS as a dependent variable, while age, education level, gender, disease duration and lesion load (i.e., the total volume of the lesions) were considered as predictors. Multicollinearity was assessed through the variance inflation factor (VIF).^50,51^ Given the relatively low numerosity of our sample, we validated our model using the leave-one-out cross validation (LOOCV). Hence, we built *n* multilinear model (where *n* is the size of the sample), each time excluding a different subject from the training set, and then verifying the ability of the model to predict the clinical disability of the excluded subject.

## Results

### Empirical Spectral Differences between patients and controls

The PSDs were estimated using the Welch method for each participant, in order to identify group differences in spectral features. In particular, the analysis focused on the alpha band (8-13 Hz). PSD estimates were averaged across ROIs to obtain a median spectrum for each participant (Fig 2 A). In line with earlier studies, we found robust differences in alpha features between MS patients and age-matched controls. A modest slowing of the peak alpha frequency was also observed upon visual inspection, however, this did not survive formal statistical testing (Fig 2, B). Most strikingly, MS patients displayed significantly attenuated peak alpha amplitudes as compared to controls (*P < 0.01*) (Fig 2 C). Furthermore, the total power in the alpha band, measured as the area under the PSD curve between 8-13 Hz, was significantly lower in MS patients *(P = 0.02* Fig 2 D). Visual inspection also indicated modest differences in the theta and beta amplitude (not shown), however these were not investigated further given the scope of this manuscript.

**Figure 1.**
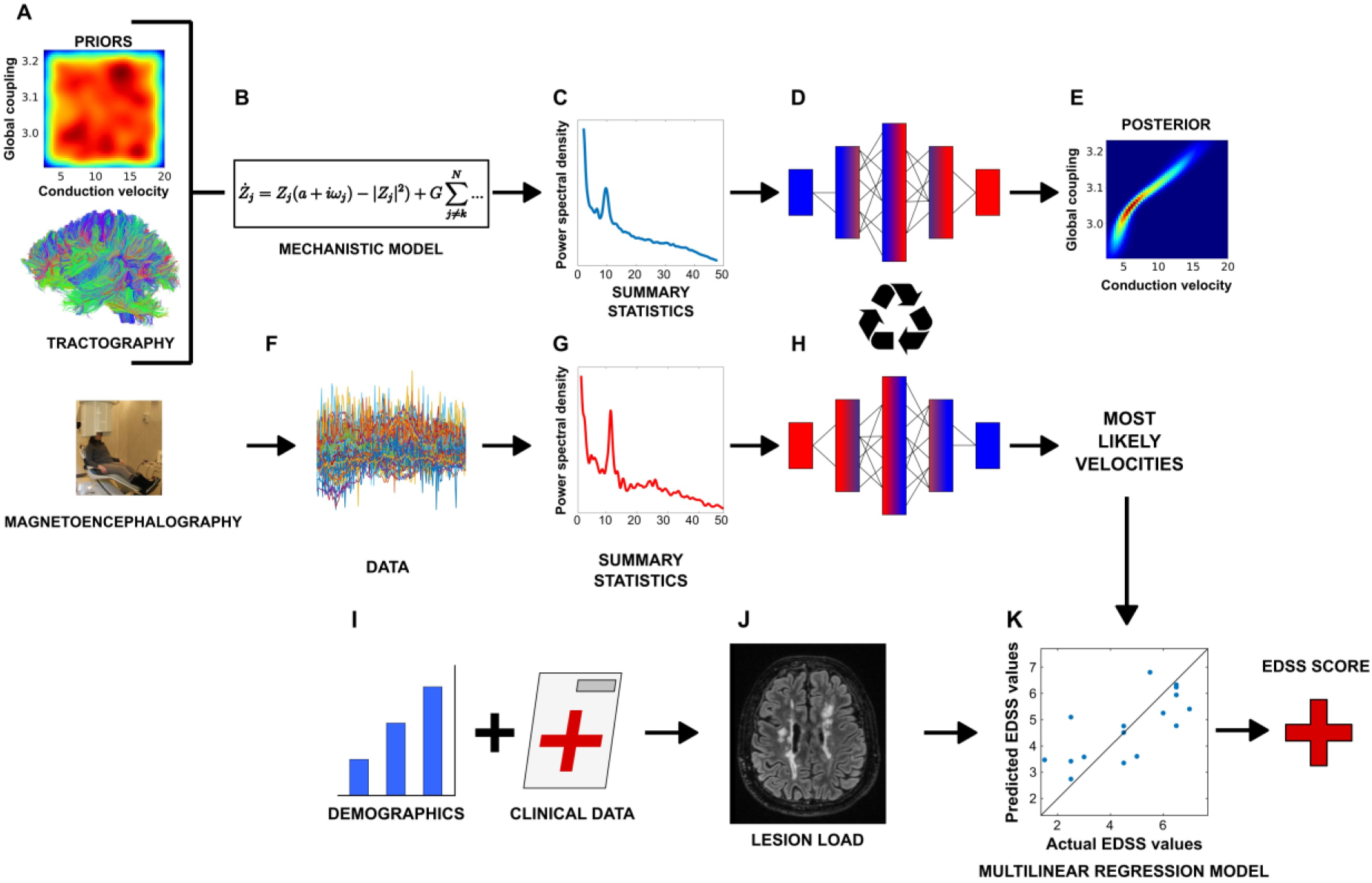
Overall pipeline. **A - B.** A subject-specific tractography was used to generate synthetic data based on global couplings and conductions velocities that were drawn randomly within physiological plausible ranges. **C.** Global spectral features were extracted from the synthetic data. **D.** Machine learning was used to learn an invertible function that would relate velocities and global coupling to spectral features. **E.** Posterior distributions of global coupling and conduction velocities. **F-G.** Spectral-properties of source-reconstructed magnetoencephalography data were extracted for MS patients and controls. **H.** The invertible function is reversed, as to estimate the most likely velocities given the observed summary statistics (i.e. spectral features). **I.** Demographic features and clinical data. **J.** The picture shows an example of the magnetic resonance of a patient with MS, showing the typical lesions. The total volume of the lesions is included, along with clinical data and demographics, in **K.** A multilinear model that predicts the individual clinical disability, as measured by the EDSS.

**Figure 2.**
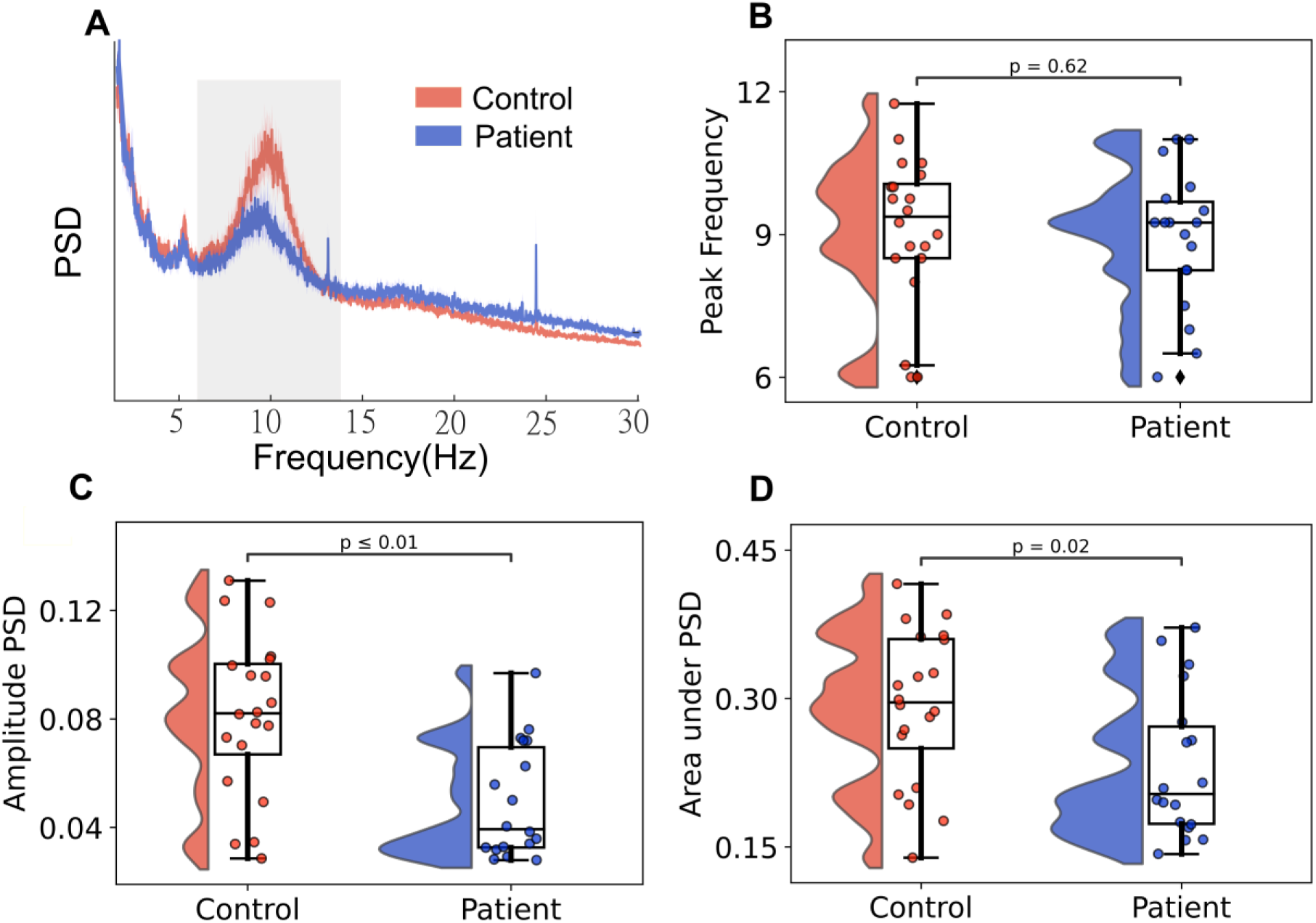
Empirical data features. Data features extracted from empirical MEG data for control and MS groups. **A.** Median power spectral density plotted for Controls (red) and MS patients (blue) in the 1-30 Hz range. Shaded region represents the alpha band (8-13 Hz). **B.** The peak frequency shows non-significant changes in the control group relative to the patients group (*P = 0.62*). **C.** The amplitude of peak frequency illustrates a significant decrease from the control to the patients group (*P < 0.01*). **D.** The area under the PSD shows a significant decrease from the control to the patients group (*P = 0.02*).

### Numerical solution of the Stuart - Landau model

The complex interplay of coupling (indexed by *G*) and average conduction velocity (*V*) modulates the average peak frequency, peak power and spectral distribution of oscillators in the network (see Fig 3). As shown previously, when operating in the partially synchronized regime, oscillators organize as metastable clusters, each oscillating at an emergent frequency and amplitude.^16,17,52^ A similar frequency slowing has been previously explored for networks of phase oscillators like the Kuramoto model.^17,22^ However, the Stuart-Landau model additionally allows for amplitude coupling.^16^ In short, for weak couplings and large delays, the network resists synchronization and oscillators drift close to their natural frequencies (40 Hz). This state is characterized by relatively high gamma amplitudes, since all the power of the system is confined in the gamma band, that is the natural frequency of the oscillators. In contrast, the network synchronizes at frequencies lower than the natural frequency (*ω* < *ω*_0_) for stronger couplings, due to conduction delays.^17^ Generally, for a given average conduction speed, stronger coupling is associated with the emergence of slower and lower amplitude oscillations. As one can observe in Fig 3, panels A and B, traversing leftwards along the heatmaps (that is, assuming progressively slower velocities or, equivalently, increasing delays) yields diminished peak frequency and power, provided the delays remain below the synchronization threshold.

**Figure 3.**
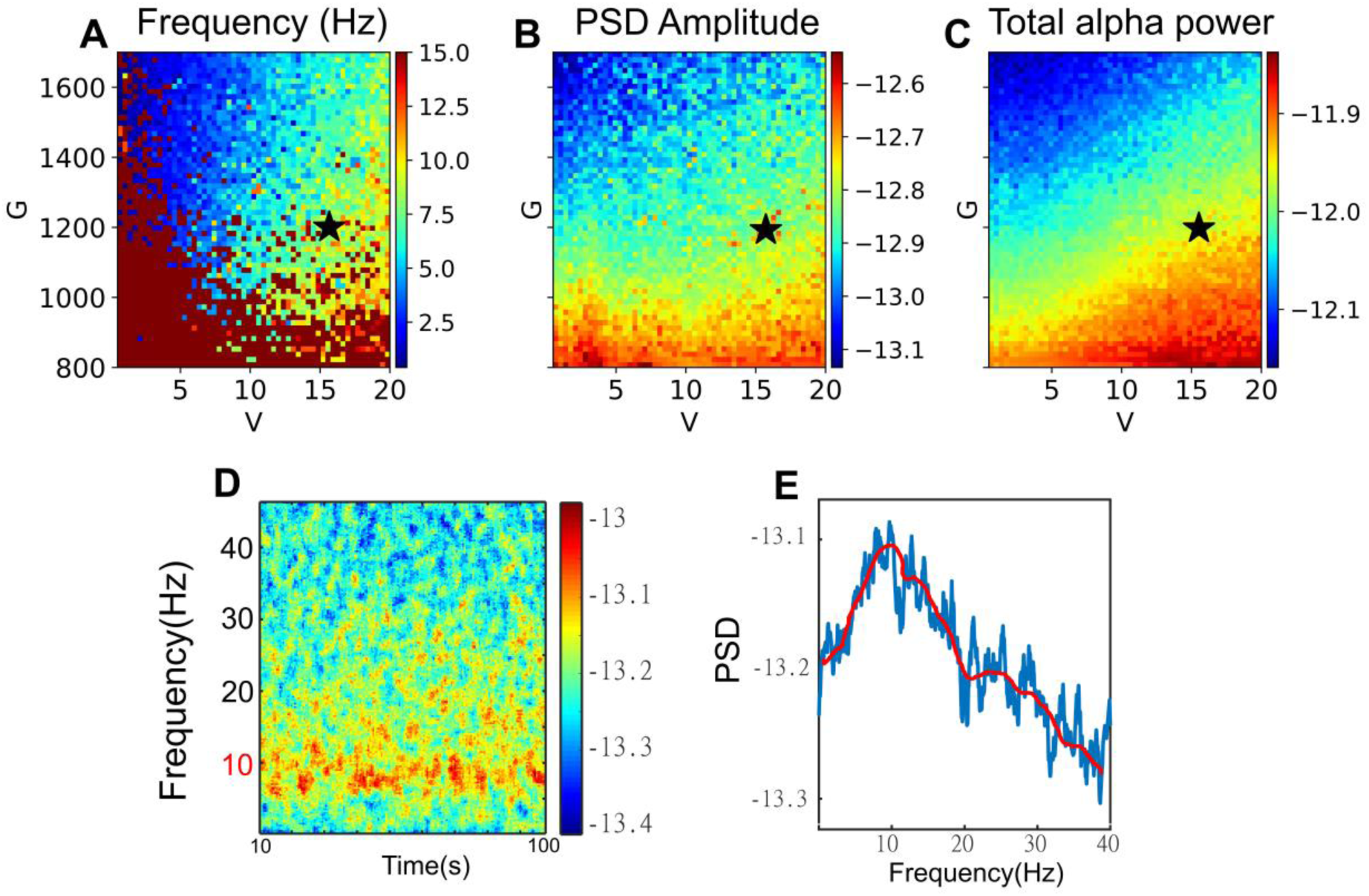
Model output. **A.-C.** Heatmaps indicate the median frequency, the median peak alpha PSD and the total alpha power, respectively, for the range *G* ∈ [800, 1800] and *V* ∈ [1, 20]. **D.** Simulation output represented as the median spectrogram for *G*=1200, *V*=16 m/s (indicated as * in heatmaps) and **E.** The corresponding PSD, showing a peak in the alpha band.

### Model inversion and delay inference

Forward models predict the effects knowing the causes, whereas inverse models reveal the causes given the effects.^53^ In this work, model inversion is used to estimate the most likely conduction velocities (and global coupling, i.e., the causes) in any individual given their spectral features (i.e., the effects). To obtain such estimation, we used a mechanistic personalized generative brain model, where one can simulate the effect of changing conduction velocities on the spectral features of individual source-reconstructed MEG signals (via well-studied changes in network synchronization).^17^ Simulating the effects of different conduction velocities allows the estimation of their posterior distributions (i.e., what is the probability of a particular delay given the observed spectral features?). In this study, we utilized simulation-based inference (SBI) to perform Bayesian model inversion, which provides us with an efficient estimation of the posterior distributions of the conduction velocities (and global coupling). This results from updating the prior probability of having a plausible range of conduction velocities with the observed data (i.e., the spectral features), through the likelihood function, which in our case relies on personalized, mechanistic, large-scale brain models, and well-defined physical properties of networks of weakly coupled oscillators. In particular, we used ANNs for probability density estimation (i.e., Masked Autoregressive Flow) trained with a budget of 20k model simulations.

Fig 4 shows the estimated posterior distributions over the whole-brain model parameters (the global coupling strength *G*, and the velocity *V*), by using SBI against the empirical PSD, pooled over the control and the patient groups. The posterior distribution of the parameter *G*, which scales the structural connectivity of subjects, demonstrated non-significant changes (p-value = 0.87), whereas the posterior distribution of averaged velocities *V* significantly decreased (p-value <0.01) in MS patients as compared to the control group.

**Figure 4.**
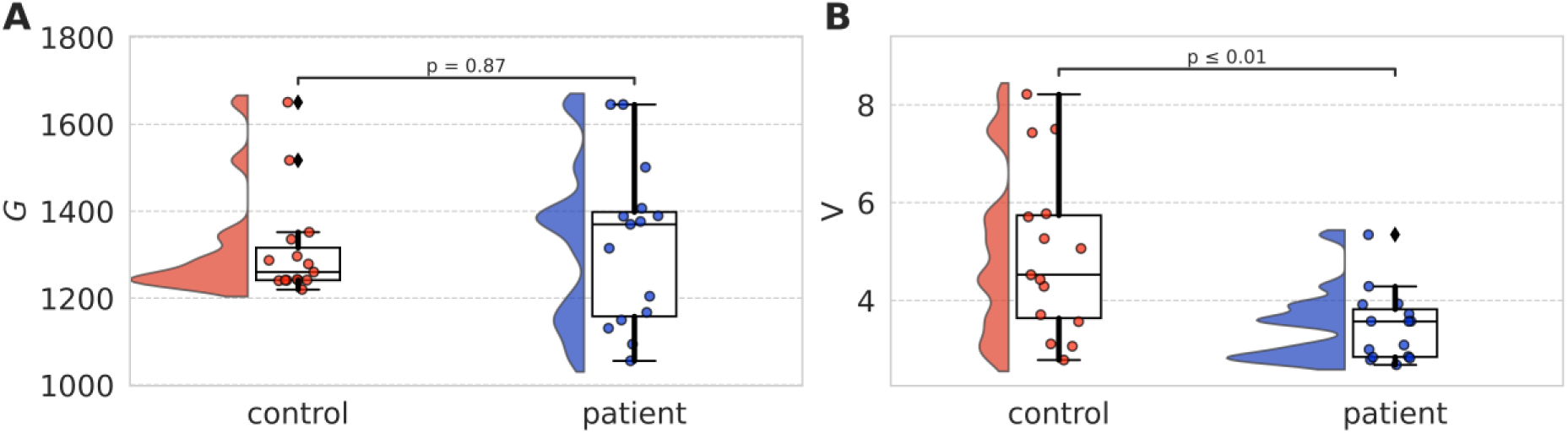
Estimated parameters. Estimated posterior distribution for two brain parameters from the PSD of the empirical MEG data, pooled over control and MS patient groups. **A.** The global coupling strength *G* shows non-significant changes (*P = 0.87*). **B.** The velocity parameter *V* significantly decreases in the control group relative to the patient group (*P < 0.01*).

Using synthetic data for model inversion allowed us to validate our inference process by knowing the ground-truth of the parameters being estimated (as demonstrated in Fig 5). Firstly, it can be seen that the predicted PSDs closely resemble the empirical ones (Fig 5, panel A, blue and red, respectively). An example of observed and predicted MEG time-series averaged across brain regions are shown in Fig 5 panel B (by transforming the observation from the frequency-domain to the time-domain). Our result indicates that SBI accurately estimates a posterior distribution that narrows around the true parameters which generated the observed data (see Fig 5, panels C and D). This confirms that SBI can accurately capture relevant PSD features of MEG data, including the peak, amplitude and total power of the PSDs.

We also checked the accuracy of the estimated posterior distributions, in that increasing the number of simulations from 100 to 20k led to progressively tighter posteriors (i.e., less uncertainty in the estimation, as shown in Fig 5, panels C and D), with the maximum a posteriori closer to the ground-truth (i.e., the parameters used to generate the data). The joint posterior distribution (i.e., on the same probability space) between parameters *G* and *V* indicates a high correlation of 0.75 (Fig. 5, panel E). In terms of computational cost, each model simulation took around 12 sec (using just-in-time compilation in Python), which can be easily run on multiple CPU/GPU cores. Using a MAF, the training took around 3 mins, whereas generating 10000 samples from the posterior took only 1 sec. We then performed sensitivity analysis using the estimated posterior from observation, which indicated that the model is more sensitive to the parameter *V* than *G*, as conveyed by the larger eigenvalue (by around 7 orders of magnitude), which reflects a larger gradient of the posterior for the averaged velocity (see Fig 5, panel F).

**Figure 5.**
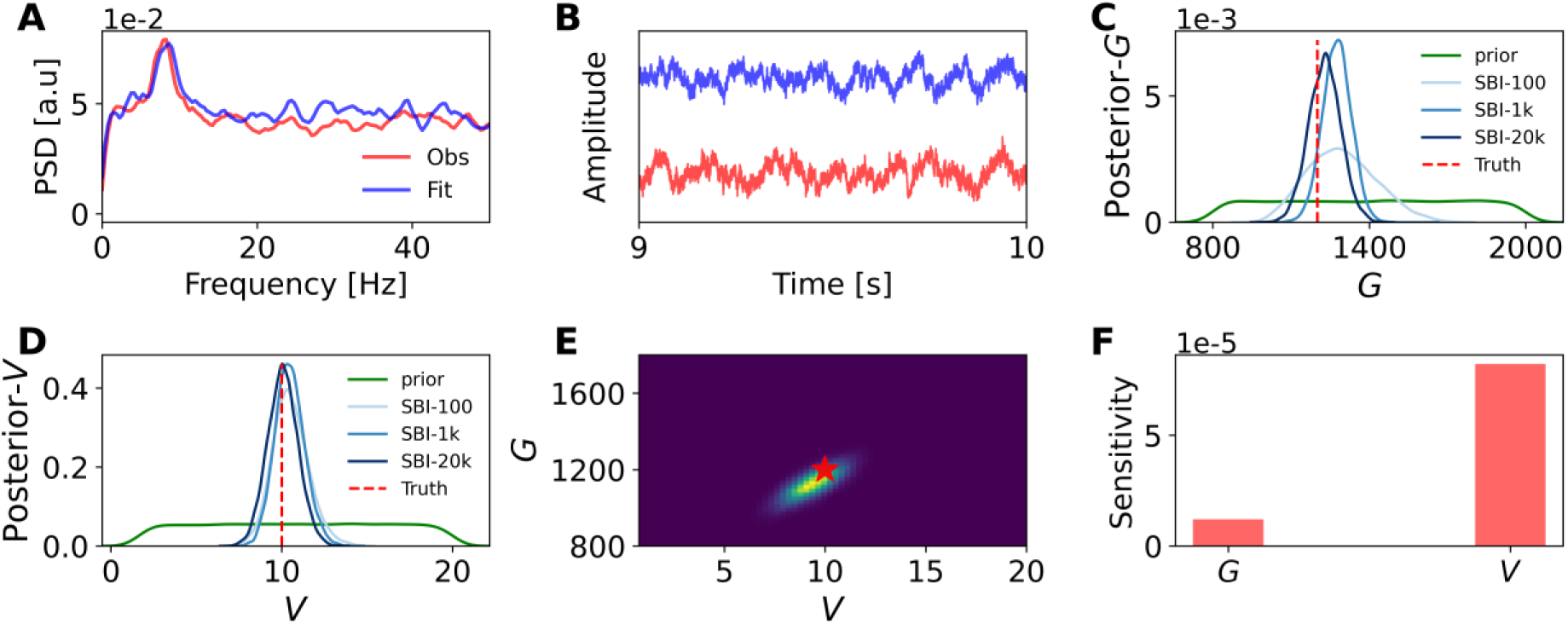
Diagnostics of the inference process. **A-B.** Observed (red) and predicted (blue) PSD of MEG data and corresponding time-series averaged over brain regions, respectively. **C-D.** Inferred posterior distributions for the global coupling strength *G*, and the velocity *V*, respectively, given PSD features (amplitude, median frequency and total power). Increasing the number of simulations for training steps yields progressively tighter posteriors and, thus, a more accurate estimate. **E.** Joint posterior distribution between parameters *G* and *V* estimated from 20k simulations (correlation=0.75). The ground truth parameters are shown in red, the high-probability parameters in yellow, the low-probability ones in blue. **F.** Sensitivity analysis using the estimated posterior, indicating stronger model sensitivity to *V* than to *G* (the Eigenvalues for *G* and *V* are 1.2e-05, and 8.2e-05, respectively).

These results are consistent by normalizing the parameters *G* and *V* between zero and one (see Fig S1, in supplementary materials). In sum, our findings indicate that SBI on PSD at whole-brain level is efficient and accurate for Bayesian model inversion of MEG data.

### Prediction of clinical outcome

We then moved on to test the validity of the inferred individual delays in terms of prediction of the clinical disability. Please note that the EDSS score was not available for one patient and, hence, the prediction of the clinical outcome was carried out on 17 patients only. To this end, we built a multilinear model to predict clinical disability as measured by the EDSS scale. Gender, age, education level, disease duration and lesion load (i.e., the total volume of lesions), and the inferred conduction velocities were considered as predictors. We found that the model performs well at predicting individual disability (R2 = 0.595, Adjusted R2 = 0.41, see Fig 6, panel A). Adding the inferred velocities to the model significantly improves the predictive power over the EDSS (*P* =0.028). The predicted vs. empirical EDSS values are shown in Fig 6, panel B. To test for the generalizability of our model, we used a leave-one-out cross validation (LOOCV) scheme, whereby the model is trained, at each iteration, by excluding one patient, and then the EDSS is predicted for the patient that was not included in the training data. The results of the cross-validation are shown in the lower row of Fig 6 (i.e., the average results across all iterations are reported). In this case, the average R2 is 0.61, adj R2 = 0.40), confirming the predictive power of the model. Again, adding the estimated speed to the model significantly improved the predictive power (*P = 0.0417*). Panels C and F show the distribution of the residuals, confirming the appropriateness of using a linear model in this context. Finally, the values of Variance Inflation Factor (VIF) were always below two for all predictors, showing that multicollinearity is unlikely to affect the model (not shown). The model remains predictive of individual disability also when the power spectral density in the alpha range is added as a covariate (supplementary material 2).

**Figure 6.**
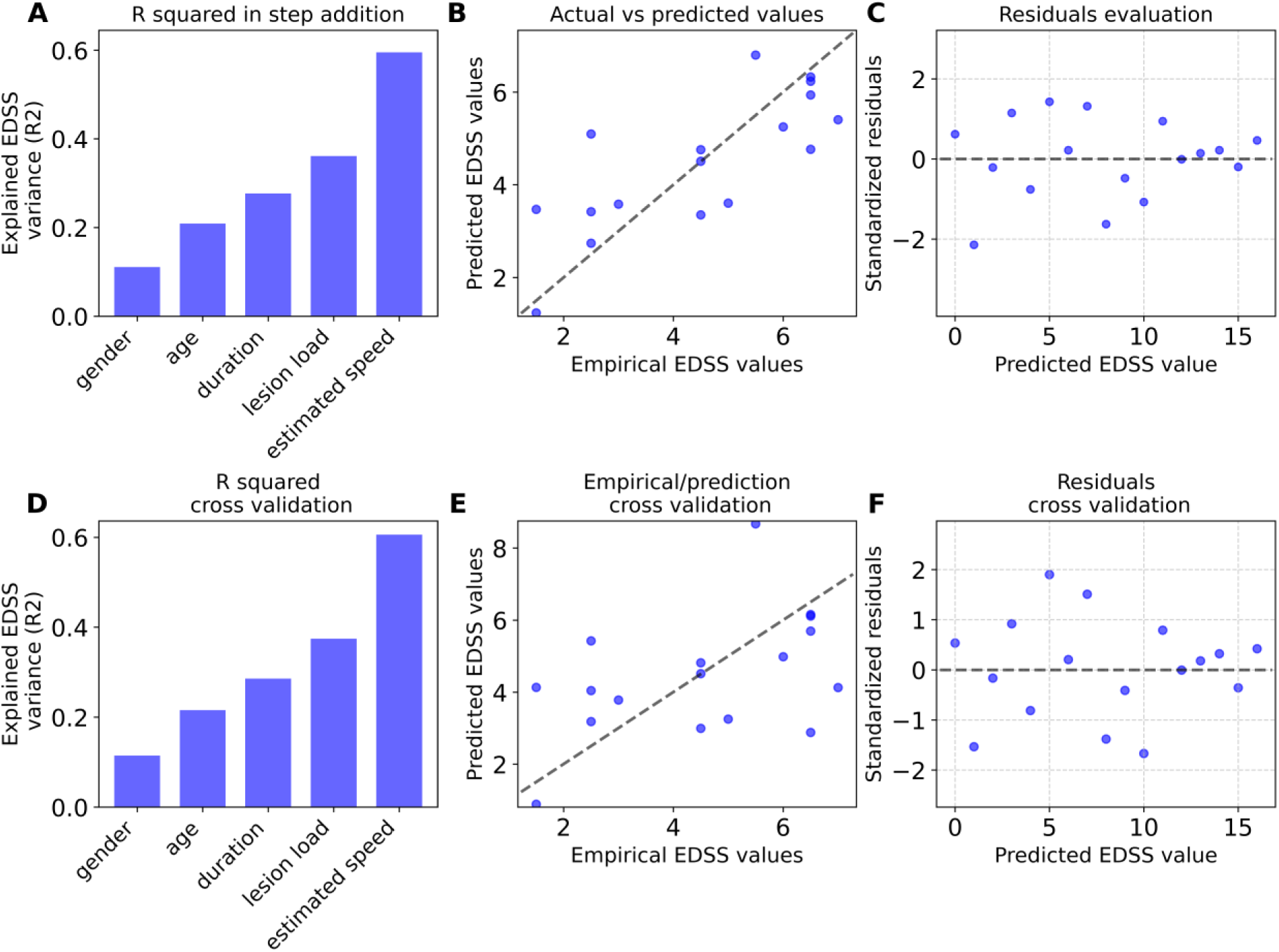
Prediction of clinical outcome. Multilinear regression model with leave-one-out cross validation (LOOCV) performed to test the capacity of the estimated speed to predict the EDSS scores in MS patients. **A., D**. Variance explained by the additive model including five variables (i.e., gender, age, disease duration, lesion load and estimated speed). Adding the estimated conduction velocities significantly increased the predictive power in both classical multilinear (panel **A**, *P = 0.028*) and cross-validated (panel **D**, *P = 0.0417*) models. **B., E.** Empirical and predicted EDSS scores. **C., F.** standardized residuals of the model.

## Discussion

In this manuscript we set out to test two related hypotheses. First, we hypothesized that slower conduction velocities across the brain manifest themselves as a reduction of the alpha power and a shift in the alpha peak at the level of the whole-brain frequency spectrum. Second, we hypothesized that the augmented delays are predictive of the clinical disability in MS, above and beyond the clinical damage as measured by the lesion load. To test our hypotheses, we used source-reconstructed MEG data and tractographies from 18 MS patients and 20 healthy controls.^12^ Most specifically, the area under the PSD in the alpha band, as well as the peak in the alpha band, are both lower in MS patients as compared to controls. This allowed us to hypothesize a direct link between the large-scale frequency spectrum and some well-known properties of weakly-coupled oscillators, whereby the overall frequency of oscillation of a network is expected to decrease (with respect to the natural frequency of the oscillators) with growing delays.^17,22^ With this in mind, we used a realistic, mechanistic, mean-field based model of neuronal ensembles, namely Stuart-Landau oscillators,^44^ coupled according to the subject-specific connectomes. We could then successfully reproduce in silico, for each individual, the expected shift in the peak spectral amplitude and the total power as a function of growing delays. This provides a mechanistic account of the supposed link between the modifications in the individual large-scale frequency spectrum (i.e., our observed quantity) and the conduction delays (which cannot be observed directly in MS at the whole-brain level). The presence of such a mechanistic link allowed us to use this model to infer the individual average delays, in the sense of performing Bayesian simulation-based inference in order to estimate the most likely individual delays, given the observed subject-specific large-scale frequency changes. Crucially, our large-scale model does not include any informative priors about the delays (we used a uniform distribution in a biophysically plausible range). Despite this, the results of the model inversion clearly point to the fact that slower velocities (i.e., longer delays) become more likely given the observed changes in the power spectra in MS patients. In fact, the estimated velocities in MS patients are significantly slower than the ones estimated in the controls.

Subsequently we used the inferred individual velocities to predict individual clinical impairment. In accordance with our hypothesis, the predictive power of our model significantly increases when we include the inferred velocities in the model. The lesion load was also included as a covariate. However, lesion load alone failed to predict most of the variance in clinical disability, in accordance with previous evidence.^3–5^ We propose that functional delays would be a relevant feature to explain clinical disability in MS. In that sense, we would interpret the poor performance of structural lesions at predicting clinical disability as the consequence of the fact that it is the delays and not the structural lesion per se to be causing clinical disability. However, functional delays depend on a multitude of different elements, including the size and the myelination of individual structural tracts, as well as the network in which each tract is embedded.^12^ As such, it is evident that structural lesions would be a proxy of the functional damage that can only partly convey the ‘true’ functional damage. In accordance with this view, the inferred functional delays drastically outperform the lesion load at predicting clinical impairment. In this sense, our results suggest a principled, physics-informed way to directly predict individual disability from the inferred delays, thereby obtaining the most clinically relevant large-scale feature in MS by directly inferring the individual functional effects of the lesions instead.

Furthermore, given that the lesion pattern is patient-specific in MS, the way edges are affected is most likely non-homogeneous even within each individual patient. However, in order to make the Bayesian inference computationally feasible, we adopted the simplifying assumption of homogeneous slowing of velocities across the brain of each-patient. This assumption might be reasonable given the recent evidence showing that the slowing of velocities across the brain of MS patients is not limited to the lesioned edges but, rather, it is widespread across the brain^12^. However, further improvement might be obtained including information about the individual lesions as model priors, as to introduce yet one more level of personalization in the large-scale models. This is feasible due to the flexibility of the Bayesian approach used in this study as compared to the classical optimization methods, which allows us to integrate the prior information in the inference process. For instance, it has been shown that measuring the prediction accuracy of a whole-brain network model with a higher level of information encoded in priors provides decisive evidence in favor of the true hypothesis regarding the degree of epileptogenicity across different brain regions.^54,55^ To estimate the model parameters, optimization methods (within the Frequentist approach) have been used in previous studies^16,56^ by defining an objective (or a cost) function to score the performance of the model against the observed data. However, such a parametric approach results in only a point estimation, and the optimization algorithms may easily get stuck in a local minimum, requiring multi-start strategies to address the potential multi-modalities. Moreover, the estimation depends critically on the form of the objective function defined for optimization. These issues can be solved by using Bayesian SBI. On the one hand, the traditional approaches for SBI^57^ suffer from the curse of dimensionality and are sensitive to the ad-hoc choices (i.e., rejection thresholds, distance functions, and summary statistics), which significantly affects both the computational efficiency and the accuracy; on the other hand, the SBI with the deep neural density estimator used in this study transforms a simple distribution conditioned on the data features to obtain the full probability distribution of the target parameter, while dealing with non-linear and high-dimensional latent spaces and highly structured data (without the sensitivity to the tolerance level in the accepted/rejected parameter setting).

Importantly, while promising, our results need to be replicated in larger multimodal, multicentric cohorts, to check that they are not specific to this particular dataset. Although it will be important to check the feasibility of using EEG instead of MEG, as this would contribute to the widespread applicability of our approach, the pipeline we propose (in terms of the building of a subject-specific, large-scale brain model and the Bayesian model inversion to infer the individual delays) is available using the virtual brain (https://www.thevirtualbrain.org), a free, python-based tool designed for personalized large-scale models.

In conclusion, we propose a principled way to link the most relevant physiopathological feature in MS, namely the slower conduction velocities across the brain network induced by myelin damage, to a large-scale observable quantity, that is the modification of the frequency spectrum. We then merged structural resonance, source-reconstructed MEG data, and a mechanistic model of the large-scale brain with using state-of-the-art deep learning algorithms, in order to infer the subject-specific median conduction velocities. Finally, we use the inferred conduction velocities in order to predict individual disability. Thus, our work paves the way to a principled application of personalized large-scale models in MS. Further, from a more pathophysiological viewpoint, our results shed light on the known, poor correlation between lesion load and clinical disability-the “clinico-radiological paradox”, by suggesting and then quantifying the potential mechanisms (i.e., lower conduction velocities) through which structural damage would provoke large-scale changes in brain activity and, in turn, clinical impairment.

## Author contribution

Pierpaolo Sorrentino, Anagh Pathak and Meysam Hashemi: conceptualization; Pierpaolo Sorrentino, Anagh Pathak, Abolfazl Ziaeemehr and Emahnuel Troisi Lopez: methodology development; Pierpaolo Sorrentino, Emahnuel Trosi Lopez, Lorenzo Cipriano and Antonella Romano: Investigation; Pierpaolo Sorrentino, Emahnuel Troisi Lopez and Ablofazl Ziaeemehr: Visualization; Arpan Banerjee, Giuseppe Sorrentino, Meysam Hashemi and Viktor Jirsa: supervision; Pierpaolo Sorrentino, Emahnuel Troisi Lopez, Lorenzo Cipriano, Antonella Romano, Maddalena Sparaco and Mario Quarantelli: data collection and curation; Pierpaolo Sorrentino, Emahnuel Troisi Lopez, Lorenzo Cipriano, Antonella Romano, and Mario Quarantelli: data processing; Pierpaolo Sorrentino, Anagh Pathak and Meysam Hashemi: writing original draft; all authors: writing—review & editing.

## Data availability

The MEG data and the reconstructed avalanches are available upon request to the corresponding author (Pierpaolo Sorrentino), conditional on appropriate ethics approval at the local site. In case data are requested, the corresponding author will request an amendment to the local ethical committee. Conditional to approval, the data will be made available. The Matlab code is available at https://github.com/Ziaeemehr/virtual_multiple_sclerosis_patient

