## Supplementary materials for "The virtual multiple sclerosis patient: on the clinical-radiological paradox"

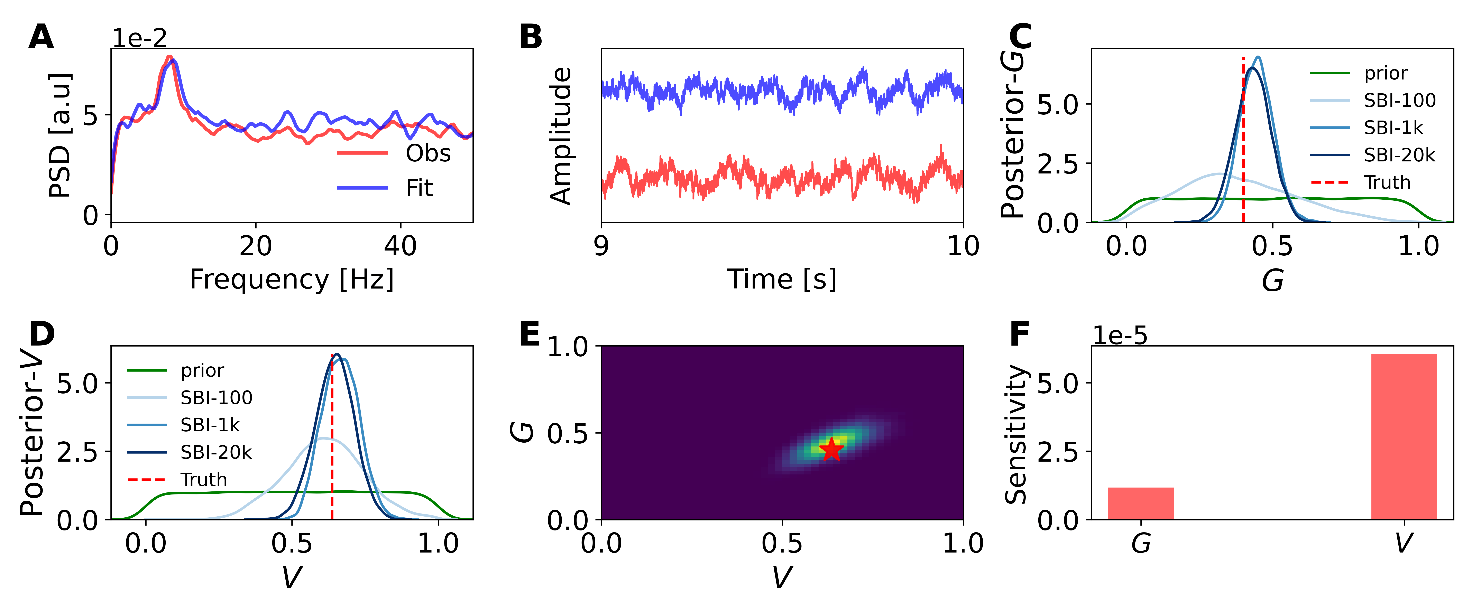

**Fig S1 Diagnostics of the interference process with normalized parameters.** The results of SBI are consistent by normalizing the parameters between zero and one.  **A-B.** Observed (red) and predicted (blue) PSD of MEG data and corresponding time-series averaged over brain regions, respectively.  **C-D.** Inferred posterior distributions for the global coupling strength G, and the velocity V, respectively, given PSD features (amplitude, median frequency and total power). Increasing the number of simulations for training steps yields progressively tighter posteriors and, thus, a more accurate estimate. **E.** Joint posterior distribution between parameters G and V estimated from 20k simulations (correlation=0.67). The ground truth parameters are shown in red, the high-probability parameters in yellow, the low-probability ones in blue.  **F.** Sensitivity analysis using the estimated posterior, indicating stronger model sensitivity to V than to G (the Eigenvalues for G and V are 1.17e-05, and 6.4e-05, respectively).

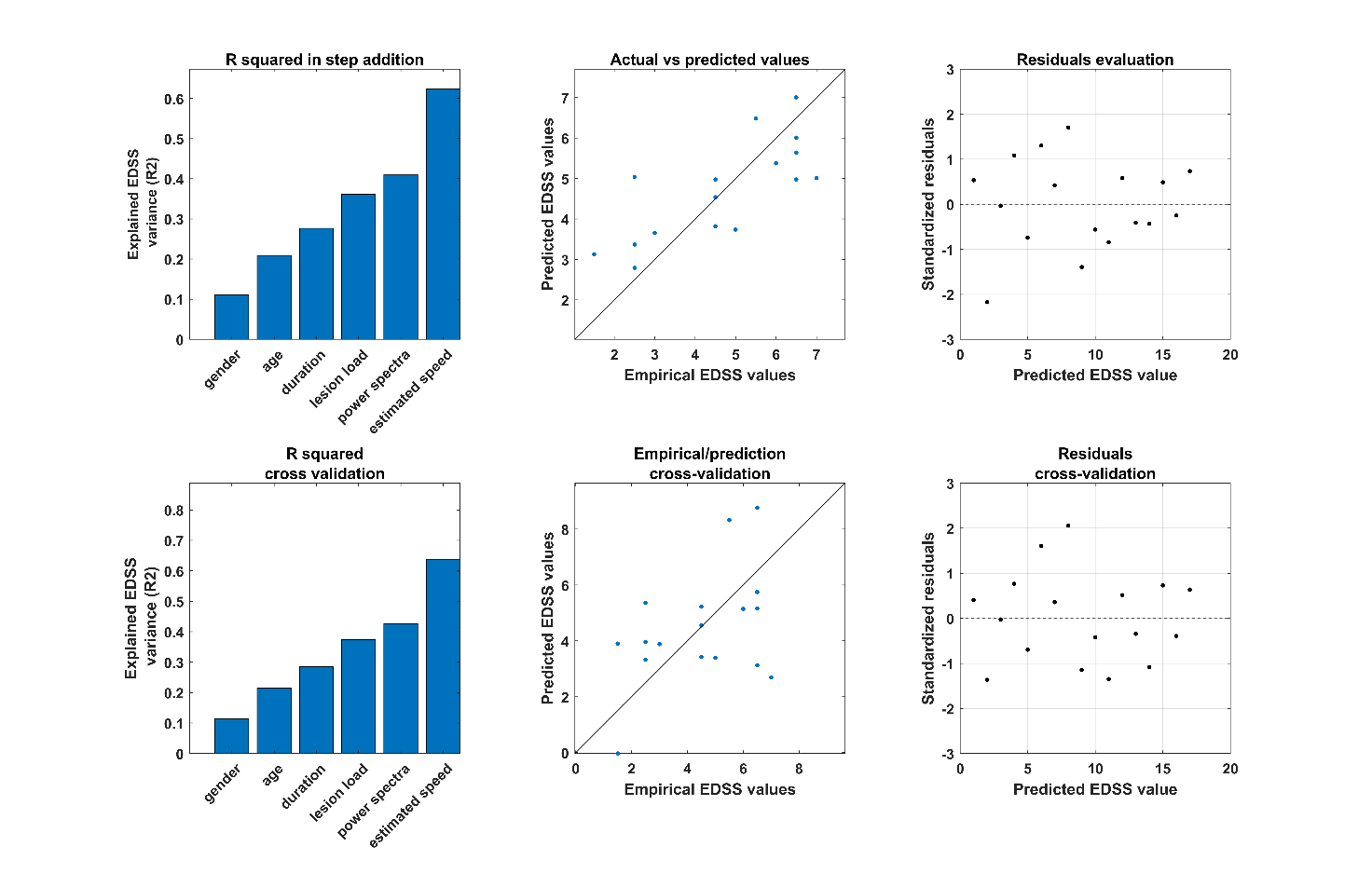

**Fig S2. Prediction of clinical outcome including power spectra.**  Results of the behavioural model when the area under the curve is added as a covariate.
